# Estimating the sensitivity of non-treponemal and treponemal antibody tests in primary syphilis

**DOI:** 10.1101/2025.11.07.25339586

**Authors:** Stephanie F. Sweitzer, Jane S. Chen, Mitch M. Matoga, Ligang Yang, Eduardo Lopez-Medina, Jonny A. Garcia-Luna, Irving F. Hoffman, Bin Yang, Juan C. Salazar, Justin D. Radolf, Jonathan B. Parr, Arlene C. Seña

**Affiliations:** Division of Infectious Diseases, Department of Internal Medicine, University of North Carolina, Chapel Hill, North Carolina, USA; UNC Project Malawi, Lilongwe, Malawi; Dermatology Hospital, Southern Medical University, Guangzhou, China; Centro Internacional de Entrenamiento e Investigaciones Medicas – CIDEIM, Cali, Colombia; Department of Pediatrics, Universidad del Valle, Cali, Colombia; UConn Health, Farmington, Connecticut, USA; Universidad Icesi, Cali, Colombia; Connecticut Children’s, Hartford, Connecticut, USA

**Keywords:** Syphilis, diagnostics, non-treponemal testing

## Abstract

Among patients with primary syphilis, sensitivity of TRUST was 55% (6/11) compared to darkfield microscopy (DFM) and 60% (9/15) compared to *Treponema pallidum* PCR. Sensitivity of RPR was 78% (38/49) and 93% (43/46), respectively. TPPA had a sensitivity of 95% (41/43) compared to DFM and 96% (43/45) compared to PCR.

**Short Summary:** Compared to direct detection methods for the diagnosis of primary syphilis, TRUST has low sensitivity, RPR has moderate to good sensitivity, and TPPA has high sensitivity.

## Introduction

Early identification and treatment of syphilis are essential for reducing sequelae and disease transmission. Darkfield microscopy (DFM) has been utilized to detect *Treponema pallidum subsp. pallidum (Tp)* from lesions, but requires skilled technicians.^1^ Nucleic acid amplification tests, such as *Tp* quantitative polymerase chain reaction (qPCR), are highly sensitive in primary syphilis (PS),^2,3^ but are not widely available. Consequently, the diagnosis of PS has relied on serological testing, although detection of non-treponemal and/or treponemal antibodies can be delayed after initial infection.^4^ While prior studies have explored the sensitivity of non-treponemal (also referred to as lipoidal antigen) and treponemal tests for PS, more data are needed to refine sensitivity estimates.^5^

We performed a retrospective secondary analysis of data and biological specimens from participants with PS in a prior study^6^ to describe the clinical and laboratory characteristics associated with PS and to assess the sensitivity of serologic testing compared to DFM and *Tp* qPCR for PS.

## Methods

### Study setting and population

From November 28, 2019 to May 27, 2022, participants ≥18 years old with early syphilis were recruited and enrolled at three clinical sites in Guangzhou, China; Cali, Colombia; and Lilongwe, Malawi. Procedures for the parent study have been previously reported;^6^ for the present analysis, we focused only on participants who were enrolled with suspected PS.

Syphilis staging for each participant was determined by trained study staff at each site. Participants were enrolled if they had PS defined as the presence of one or more painless anogenital ulcerative lesions (e.g., chancres) and positive DFM and/or reactive non-treponemal or rapid treponemal antibody tests. Individuals were excluded if they had received antibiotics active against *Tp* within 30 days of enrollment. The study was approved by the institutional review boards (IRBs) at the University of North Carolina at Chapel Hill (IRB protocol number 19-0311), the Dermatology Hospital of Southern Medical University in Guangzhou (IRB protocol Guangdong Dermatology Hospital Lunli Shencha-20181202 [R3]), CIDEIM in Cali (IRB protocol 1289), and the National Health Sciences Research Committee Ministry of Health in Lilongwe (IRB approval 2252).^6^

Following written informed consent, signs and symptoms were recorded from study participants including the number and anatomic location of anogenital ulcer(s). Laboratory tests for syphilis included DFM from lesion swabs and rapid treponemal antibody testing (SD Bioline Syphilis 3.0, Abbott Laboratories, IL). Non-treponemal testing was performed on all participants. *Treponema pallidum* particle agglutination (TPPA) testing was conducted at sites where rapid treponemal testing was not available. Rapid testing was performed for HIV. Following DNA extraction from anogenital lesion swabs, qPCR targeting the *Tp* polymerase I gene (*polA*/*tp0105*) was performed.^7^ Any sample with >0 copy numbers was considered *Tp* positive. All participant data were collected in case report forms uploaded into a secure Research Electronic Data Capture database.

### Statistical analysis

Categorical variables were summarized using frequency counts and percentages. Continuous variables were summarized by medians and interquartile ranges. Sensitivities were calculated with 95% confidence intervals using the binomial distribution. All analyses were performed with Stata version 13.1 (StataCorp, College Station, TX).

## Results

Over the study period, 2,820 individuals were screened and 79 participants were enrolled with PS. Median participant age was 27 years, with an interquartile range (IQR) of 23-32 years (Table 1). The majority were male (61, 77%) and most reported their sexual orientation as heterosexual (62, 78%). Most identified as Black African (44, 56%), with fewer identifying as Asian (18, 23%) or Hispanic/Latino (17, 22%). Nine (11%) reported a prior history of syphilis or other genital ulcer disease. Twelve (15%) reported that they were living with HIV, and of those, 8 (67%) were on antiretroviral therapy.

**Table 1.** Characteristics of Participants with Primary Syphilis.

| **Table 1. Characteristics of Participants with Primary Syphilis.** |  |  |
| --- | --- | --- |
|  | **n (N=79)** | **%** |
| **Demographic characteristics** |  |  |
| Age in years (median, IQR) | 27 | 23-32 |
| **Sex** |  |  |
| Male | 61 | 77 |
| Female | 18 | 23 |
| **Sexual orientation** |  |  |
| Heterosexual | 62 | 78 |
| Gay | 10 | 13 |
| Bisexual | 5 | 6 |
| Other | 1 | 1 |
| Unknown | 1 | 1 |
| **Race/ethnicity** |  |  |
| African American | 1 | 1 |
| Asian | 18 | 23 |
| Black African | 44 | 56 |
| Hispanic/Latino | 17 | 22 |
| White | 1 | 1 |
| **Clinical history** |  |  |
| History of syphilis or other genital ulcer disease | 9 | 11 |
| **History of other STIs** |  |  |
| None | 63 | 80 |
| Chlamydia | 0 | 0 |
| Gonorrhea | 6 | 8 |
| Genital herpes | 1 | 1 |
| Genital warts | 2 | 3 |
| Genital ulcer disease | 3 | 4 |
| Living with HIV | 12 | 15 |
| Currently pregnant | 2/18 | 11 |
| **Clinical findings on examination** |  |  |
| Presence of ulcer/chancre | 79 | 100 |
| Penis – 1 lesion | 31 | 39 |
| Penis – 2 or more lesions | 29 | 37 |
| Perianal – 1 lesion | 1 | 1 |
| Labia/vulva – 1 lesion | 2 | 3 |
| Labia/vulva – 2 or more lesions | 14 | 18 |
| Vagina – 1 lesion | 1 | 1 |
| Vagina – 2 or more lesions | 1 | 1 |
| Mouth* – 1 lesion | 1 | 1 |
| Mouth* – 2 or more lesions | 1 | 1 |
| Other anatomic location* – 1 lesion | 1 | 1 |
| Other anatomic location* – 2 or more lesions | 2 | 3 |
| Lymphadenopathy | 13 | 16 |
| Rash<sup>†</sup> | 1 | 1 |
| Alopecia<sup>†</sup> | 1 | 1 |
| Mucous patch<sup>†</sup> | 1 | 1 |
| Condyloma lata<sup>†</sup> | 1 | 1 |
| Number of days since ulcer/chancere first appeared (median, IQR) | 9 | 7-15 |
| **Laboratory findings** |  |  |
| Positive rapid HIV test<sup>‡</sup> | 5/71 | 7 |
| Positive HIV-1 ELISA or antibody test | 1/19 | 5 |
| Positive rapid treponemal test | 20/21 | 95 |
| Reactive non-treponemal test | 59<sup>§</sup> | 75 |
| Non-treponemal antibody test titer |  |  |
| 1:1 | 4/58 | 7 |
| 1:2 | 8/58 | 14 |
| 1:4 | 1/58 | 2 |
| 1:8 | 5/58 | 9 |
| 1:16 | 7/58 | 12 |
| 1:32 | 14/58 | 24 |
| 1:64 | 9/58 | 16 |
| 1:128 | 9/58 | 16 |
| 1:256 | 1/58 | 2 |
| Positive TPPA test | 47/49 | 96 |
| Positive darkfield microscopy | 60/74 | 81 |
| Positive PCR for *Treponema pallidum* | 61/79 | 77 |
| Positive anogenital lesion swab PCR | 60/78 | 77 |
| Positive skin biopsy PCR | 1/1 | 100 |

Abbreviations: IQR, interquartile range; PCR, polymerase chain reaction assay; STI, sexually transmitted infection; HIV, human immunodeficiency virus; RPR, rapid plasma reagin test; TRUST, toluidine red unheated serum test; ELISA, enzyme-linked immunosorbent assay.
\*All participants had at least one anogenital ulcer/chancere, but some also had one or more additional lesions at other anatomical sites.
<sup>†</sup>Four participants also had signs/symptoms of secondary syphilis.
<sup>‡</sup>Four positive rapid tests were in participants who had reported they were living with HIV.
<sup>§</sup>One participant had a missing titer.

All participants had an anogenital ulcer/chancre present on physical examination (Table 1). The number of days since appearance of the ulcer ranged from 1 to 150, with a median of 9 (IQR 7-15). Thirty-one participants (39%) had a single penile lesion, 29 (37%) had multiple penile lesions, 2 (3%) had a single labial/vulvar lesion, and 14 (18%) had multiple labial/vulvar lesions. Thirteen participants (16%) had lymphadenopathy,and 4 (5%) also had signs/symptoms suggestive of secondary syphilis.

Most participants (62, 78%) had rapid plasma reagin tests (RPRs), while the remainder (17, 22%) had toluidine red unheated serum tests (TRUST). Non-treponemal antibody tests were reactive for 59 participants (75%), with quantitative titers ranging from 1:1 to 1:256 (Table 1). Rapid treponemal antibody tests were positive in 20 of 21 tested (95%). TPPA was positive for 47 of 49 tested (96%). DFM was positive in 81% (60/74), and *Tp polA* qPCR was positive in 77% (61/79) of samples collected.

The sensitivity of the non-treponemal and treponemal tests among participants diagnosed with PS is shown in Table 2. Compared to DFM, RPR had a sensitivity of 78% (38/49, 95% confidence interval [CI] 63-88), TRUST had a sensitivity of 55% (6/11, 95% CI 23-83), and TPPA had a sensitivity of 95% (41/43, 95% CI 84-99). Compared to *Tp* qPCR, RPR had a sensitivity of 93% (43/46, 95% CI 82-99), TRUST had a sensitivity of 60% (9/15, 95% CI 32-84), and TPPA had a sensitivity of 96% (43/45, 95% CI 85-99).

**Table 2.** Sensitivity of Non-Treponemal and Treponemal Antibody Tests Among Participants with Primary Syphilis.

|  |  |  |  |
| --- | --- | --- | --- |
|  | **Number of positive tests** | **Total number tested** | **Sensitivity (95% CI)** |
| Sensitivity as compared to DFM |  |  |  |
| RPR | 38 | 49 | 78% (63-88) |
| TRUST | 6 | 11 | 55% (23-83) |
| TPPA | 41 | 43 | 95% (84-99) |
| Sensitivity as compared to PCR |  |  |  |
| RPR | 43 | 46 | 93% (82-99) |
| TRUST | 9 | 15 | 60% (32-84) |
| TPPA | 43 | 45 | 96% (85-99) |
| Sensitivity as compared to any direct detection method (DFM and/or PCR) |  |  |  |
| RPR | 46 | 58 | 79% (67-89) |
| TRUST | 9 | 15 | 60% (32-84) |
| TPPA | 45 | 47 | 96% (85-99) |

Abbreviations: CI, confidence interval; DFM, darkfield microscopy; RPR, rapid plasma reagin test; TRUST, toluidine red unheated serum test; TPPA, *Treponema pallidum* particle agglutination test;

We also assessed sensitivities as compared to any positive direct detection method. Among 58 participants who were positive by qPCR and/or DFM and had an RPR collected, the sensitivity of RPR was 79% (46/58, 95% CI 67-89). Among the 15 who were positive by any direct detection method and had a TRUST collected, TRUST had a sensitivity of 60% (9/15, 95% CI 32-84). TPPA had a sensitivity of 96% (45/47, 95% CI 85-99) compared to any direct detection method.

## Discussion

In this secondary analysis of participants diagnosed with PS in a multicenter study, RPR showed moderate sensitivity compared to DFM from lesion swabs and high sensitivity compared to PCR, while TRUST demonstrated low sensitivity compared to both DFM and PCR. The TPPA demonstrated high sensitivity compared to both direct detection methods. These findings suggest that a reactive RPR is supportive for the diagnosis of PS (followed by confirmatory treponemal testing), but a negative result does not rule out PS. Furthermore, TRUST appears to have poor performance for the diagnosis of PS. TPPA, however, was highly sensitive compared to direct detection methods, and may be more useful for PS diagnosis than the non-treponemal antibody tests.

*Tp* direct detection—through DFM or nucleic acid amplification testing (NAAT)— is recognized as the gold standard of PS diagnosis.^5^ Prior studies to characterize the sensitivity of RPR compared with DFM of lesion exudate found sensitivities ranging from 48.7% to 92.7%.^8–15^ One study reported that the sensitivity of RPR was as high as 92% compared to NAAT.^16^ The sensitivity of TRUST in PS has been previously found to range from 77% to 86%.^17^ TPPA is estimated to have a sensitivity of 86.2% to 100% in PS based on previous studies.^5,18^

Our findings are thus consistent with those of prior studies with regards to the sensitivity of RPR and TPPA in PS, but we found a much lower sensitivity for TRUST in PS. A chancre may appear as early as 10 days (median 21 days) after exposure to *Tp;*^19^ however, non-treponemal tests might not yet be reactive in patients presenting with PS although treponemal antibodies can be detectable as early as 2 weeks after exposure^4^. Therefore, using DFM as a comparator could skew results toward lower sensitivities for non-treponemal tests given that patients with early lesions may have high bacterial burdens but may not yet be seropositive. NAATs are generally considered to be a more sensitive detection method than DFM,^20^ given that they can detect the presence of small amounts of DNA, do not require whole *Tp* organisms, and are likely to be positive in early lesions when antibody tests may not yet be reactive.

This analysis had several limitations. First, the sample size was relatively small. In particular, very few participants—only 17—had samples collected for TRUST, limiting our analysis of this test performance. Secondly, none of the reference assays were 100% sensitive if all of the cases were true PS cases. Negative DFM and PCR results may have been due to sampling difficulties (for example, swabs may have been collected with insufficient lesion exudate), and emblematic of the limitations in currently available syphilis testing modalities. It is also possible that participants may have been incorrectly staged as PS. There was limited testing for other STIs in these clinical settings, with only one participant undergoing testing for herpes simplex virus (HSV).

Diagnosis of syphilis is notoriously challenging, and it is possible that some participants with genital ulcers of alternate etiologies (i.e. HSV, chancroid) were misclassified as having PS. However, this is unlikely given the use of direct detection methods as a comparator.

In summary, this analysis supports the use of TPPA in the diagnosis of PS, although the time required to obtain test results should be considered. This underscores the importance of using complementary tests (such as RPR and TPPA) for confirming PS diagnoses, and suggests a need for further research to identify new point-of-care assays that could be used to improve early PS diagnosis.

## Data Availability

Deidentified data produced in the present study are available upon reasonable request to the authors.

## Acknowledgements

We would like to thank Christopher M. Hennelly for his work on this project. We also wish to express our gratitude to the study participants and research staff in Guangzhou, Cali, and Lilongwe.

